# Effectiveness of COVID-19 vaccines against Omicron or Delta symptomatic infection and severe outcomes

**DOI:** 10.1101/2021.12.30.21268565

**Authors:** Sarah A. Buchan, Hannah Chung, Kevin A. Brown, Peter C. Austin, Deshayne B. Fell, Jonathan B. Gubbay, Sharifa Nasreen, Kevin L. Schwartz, Maria E. Sundaram, Mina Tadrous, Kumanan Wilson, Sarah E. Wilson, Jeffrey C. Kwong, the Canadian Immunization Research Network (CIRN) Provincial Collaborative Network (PCN) Investigators

## Abstract

**Background:** The incidence of SARS-CoV-2 infection, including among those who have received 2 doses of COVID-19 vaccines, increased substantially following the emergence of Omicron in Ontario, Canada.

**Methods:** Applying the test-negative study design to linked provincial databases, we estimated vaccine effectiveness (VE) against symptomatic infection and severe outcomes (hospitalization or death) caused by Omicron or Delta between December 6 and 26, 2021. We used multivariable logistic regression to estimate the effectiveness of 2 or 3 COVID-19 vaccine doses by time since the latest dose, compared to unvaccinated individuals.

**Results:** We included 16,087 Omicron-positive cases, 4,261 Delta-positive cases, and 114,087 test-negative controls. VE against symptomatic Delta infection declined from 89% (95%CI, 86-92%) 7-59 days after a second dose to 80% (95%CI, 74-84%) after ≥240 days, but increased to 97% (95%CI, 96-98%) ≥7 days after a third dose. VE against symptomatic Omicron infection was only 36% (95%CI, 24-45%) 7-59 days after a second dose and provided no protection after ≥180 days, but increased to 61% (95%CI, 56-65%) ≥7 days after a third dose. VE against severe outcomes was very high following a third dose for both Delta and Omicron (99% [95%CI, 98-99%] and 95% [95%CI, 87-98%], respectively).

**Conclusions:** In contrast to high levels of protection against both symptomatic infection and severe outcomes caused by Delta, our results suggest that 2 doses of COVID-19 vaccines only offer modest and short-term protection against symptomatic Omicron infection. A third dose improves protection against symptomatic infection and provides excellent protection against severe outcomes for both variants.

## INTRODUCTION

The World Health Organization declared Omicron a Variant of Concern on November 26, 2021 due to its highly transmissible nature and risk of immune evasion.^1^ In Ontario, Canada, the first detected case of Omicron was identified on November 22, 2021; within weeks, Omicron accounted for the majority of new cases. Despite very high 2-dose COVID-19 vaccine coverage (88% among those aged ≥18 years by mid-December 2021),^2^ the rate of cases among fully vaccinated individuals increased substantially during this period.^3^

A reduction in neutralizing antibodies against Omicron following second and third doses of mRNA vaccines has been established,^4-9^ but real-world data evaluating vaccine performance against Omicron infection are still emerging.^10-18^ We previously estimated vaccine effectiveness (VE) against infection (irrespective of symptoms or severity) in the initial period following identification of Omicron in Ontario, in order to support urgent public health decisions.^19^ At that time, data on symptoms were unavailable, and few cases involved hospital admission to evaluate severe outcomes. The objective of this analysis was to estimate VE against symptomatic infection and severe outcomes caused by Omicron or Delta in Ontario.

## METHODS

### Study population, setting, and design

We used the test-negative study design and linked provincial databases to estimate VE. We included symptomatic individuals aged ≥18 years with provincial health insurance who had a reverse transcription real-time polymerase chain reaction (PCR) test for SARS-CoV-2 between December 6 and 26, 2021. Our study period was selected to align with a provincial initiative to universally screen positive specimens to differentiate Omicron and Delta cases (details below) and prior to restrictions in laboratory-based PCR test eligibility announced on December 30, 2021.^20^

We excluded: long-term care residents; individuals who had received only 1 dose or 4 doses of COVID-19 vaccine(s) or who had received a second dose <7 days prior to being tested; individuals who tested positive for SARS-CoV-2 within the previous 90 days; individuals who had received 2 or 3doses of ChAdOx1 (AstraZeneca Vaxzevria, COVISHIELD) because VE for that primary schedule is known to be lower and this product has been rarely used as a third dose;^21^ those who had received non-Health Canada authorized vaccine(s) for any dose; and those who received the Janssen (Johnson & Johnson) vaccine (which, while approved for use in Canada, was largely unavailable and very rarely used [<0.1% of the Ontario population received this vaccine]^22^).

### Data sources

We linked provincial SARS-CoV-2 laboratory testing, reportable disease, COVID-19 vaccination, and health administrative databases using unique encoded identifiers and analyzed them at ICES, a not-for-profit provincial research institute (www.ices.on.ca).

### Outcomes

We identified individuals with confirmed SARS-CoV-2 infections using provincial reportable disease data and/or laboratory data.

For VE against symptomatic infection, we restricted our analysis to individuals who had at least one COVID-19-related symptom (self-reported or measured) at the time of testing, as identified in the laboratory testing data.^23^ The specimen collection date was used as the index date. Severe outcomes were defined as hospital admission or death using the earliest of the specimen collection date, hospital admission date, or death date as the index date. Due to lags in data reporting in health administrative data, we used reportable disease data to identify severe outcomes. For symptomatic individuals who tested negative for SARS-CoV-2 repeatedly during the study period and were considered controls (for both outcomes), we randomly selected one negative test.

Positive specimens identified through whole genome sequencing as B.1.1.529 lineage or found to have S-gene target failure (SGTF; a proxy measure for Omicron resulting from the amino acid 69-70 spike deletion that does not occur with Delta) were considered Omicron infections. Specimens sequenced as B.1.617 lineage or found to be negative for SGTF were considered Delta infections. Individuals with unknown or inconclusive SGTF results were excluded.^24^ Between December 6 and 24, 2021, all specimens with a positive PCR result (and a cycle threshold [Ct] value ≤35) should have been sent for testing using Thermofisher Taqpath^™^ COVID-19 PCR to identify SGTF; as such, we selected our primary study period to approximate the period of universal screening for SGTF. Prior to December 20, SGTF-positive specimens with Ct values ≤30 also underwent whole genome sequencing (WGS). In Ontario, the estimated sensitivity of SGTF relative to WGS for detecting Omicron among samples with Ct ≤30 was 98.9% and the specificity was 99.9%.^24^

### COVID-19 vaccination

To date, Ontario has primarily used 3 products (BNT162b2 [Pfizer-BioNTech Comirnaty], mRNA-1273 [Moderna Spikevax], and ChAdOx1) in its COVID-19 vaccination program. Due to fluctuating vaccine supply, varying dosing intervals and mixed vaccine schedules were employed. Using a centralized province-wide vaccine registry to identify receipt of COVID-19 vaccines, we classified individuals depending on whether they had received 2 or 3 doses of vaccine and the timing of these doses relative to the index date. We included individuals who received at least 1 mRNA vaccine for the primary 2-dose series (since a mixed schedule consisting of ChAdOx1 and an mRNA vaccine has previously been demonstrated to have similar VE as 2 mRNA vaccines).^21^ For the third dose, we included individuals who received any mRNA vaccine and also stratified our results by specific third dose product. All comparisons used those who had not yet received any doses (i.e., “unvaccinated”) by the index date as the reference group.

Third dose eligibility in Ontario began in August 2021 and expanded gradually.^25 26^ Initially, only moderately or severely immunocompromised individuals were eligible to receive a third dose as part of an extended primary series.^27^ Shortly thereafter, third doses (i.e., ‘boosters’) were provided to residents of higher-risk congregate settings for older adults (e.g., long-term care homes, high-risk retirement homes).^28^ In early October, older adults living in other congregate care settings, including all remaining retirement homes, became eligible. All individuals aged ≥70 years and healthcare workers became eligible on November 6, followed by individuals aged ≥50 years on December 13 and individuals aged ≥18 years on December 18.^29 30^ The standard interval for third dose eligibility was generally ≥168 days following a second dose but was shortened to ≥84 days on December 15.^31^

### Covariates

From various databases, we obtained information on each individual’s age (in 10-year age bands), sex, public health unit region of residence, number of SARS-CoV-2 PCR tests during the 3 months prior to December 14, 2020 (as a proxy for healthcare worker status based on the start date of the provincial COVID-19 vaccine program), past SARS-CoV-2 infection >90 days prior to index date, comorbidities associated with increased risk of severe COVID-19, influenza vaccination status during the 2019/2020 and/or 2020/2021 influenza seasons (as a proxy for health behaviours), and neighbourhood-level information on median household income, proportion of the working population employed as non-health essential workers, mean number of persons per dwelling, and proportion of the population who self-identify as a visible minority. These databases and definitions have been fully described elsewhere.^23^

### Statistical analysis

For both Omicron and Delta symptomatic infections, we calculated means (continuous variables) and frequencies (categorical variables) of baseline characteristics and compared test-positive cases and test-negative controls using standardized differences.

We used multivariable logistic regression to estimate adjusted odds ratios (aOR) comparing the odds of vaccination in each “time since latest dose” (for both second and third dose recipients) interval among cases with the odds among controls, while adjusting for all listed covariates and a categorical variable representing week of test. VE was calculated using the formula VE=(1-aOR)x100%. We estimated VE by vaccine schedule and time since latest dose.

All analyses were conducted using SAS Version 9.4 (SAS Institute Inc., Cary, NC). All tests were two-sided and used p<0.05 as the level of statistical significance.

#### Sensitivity analyses

We performed two sensitivity analyses. First, we included an additional 2 weeks of data (November 22 to December 5, 2021), representing the period immediately following Omicron identification in the province, in order to align with prior estimates of VE against infection and to observe how VE estimates against symptomatic Omicron infection changed over time.^19^ Due to the rapid rise of Omicron during the study period, we estimated VE cumulatively over time by extending the study period by 1-week increments to see how VE against Omicron changed as it became the predominant strain. We also performed a sensitivity analysis for severe outcomes using a less specific definition of Omicron to allow for a larger sample size. In this sensitivity analysis, specimens collected from December 21 onward (when the prevalence of SGTF among cases in the province was >90%) with no or inconclusive SGTF results were considered to be Omicron.

#### Supplemental analysis

We also repeated our primary analysis (i.e., December 6-26, 2021) using any infection (as opposed to symptomatic infection) in order to compare results from an updated time frame to our earlier pre-print.

## RESULTS

Between December 6 and 26, 2021, we included 16,087 Omicron-positive cases, 4,261 Delta-positive cases, and 114,087 test-negative controls. Compared to controls, Omicron cases were: younger (mean age 36.0 years vs. 42.0 years); more likely to be male; less likely to have any comorbidities; less likely to have received an influenza vaccine; more likely to have occurred during the last week of the study period; less likely to have previously tested positive for SARS-CoV-2; more likely to have received 2 doses of COVID-19 vaccines; and less likely to have received a third dose (Table 1).

**Table 1.** Descriptive characteristics of subjects tested for SARS-CoV-2 with COVID-19-relevant symptoms during the period December 6 to 26, 2021, comparing Omicron cases and Delta cases with SARS-CoV-2-negative controls

|  | **SARS-CoV-2**<br>**negative, n (%)<sup>a</sup>** | **Omicron,**<br>**n (%)<sup>a</sup>** | **SD<sup>b</sup>** | **Delta,**<br>**n (%)<sup>a</sup>** | **SD<sup>b</sup>** |
| --- | --- | --- | --- | --- | --- |
| **Total** | N=114,087 | N=16,087 |  | N=4,261 |  |
| Subject characteristics |  |  |  |  |  |
| Age (years), mean (standard deviation) | 42.0 ± 16.5 | 36.0 ± 14.1 | 0.39 | 44.2 ± 16.8 | 0.13 |
| Age group (years) |  |  |  |  |  |
| 18–29 | 30,947 (27.1%) | 6,813 (42.4%) | 0.32 | 960 (22.5%) | 0.11 |
| 30–39 | 28,387 (24.9%) | 3,467 (21.6%) | 0.08 | 941 (22.1%) | 0.07 |
| 40–49 | 19,007 (16.7%) | 2,821 (17.5%) | 0.02 | 851 (20.0%) | 0.09 |
| 50–59 | 16,695 (14.6%) | 1,922 (11.9%) | 0.08 | 679 (15.9%) | 0.04 |
| 60–69 | 11,109 (9.7%) | 723 (4.5%) | 0.21 | 450 (10.6%) | 0.03 |
| 70–79 | 5,386 (4.7%) | 233 (1.4%) | 0.19 | 269 (6.3%) | 0.07 |
| ≥80 | 2,556 (2.2%) | 108 (0.7%) | 0.13 | 111 (2.6%) | 0.02 |
| Male sex | 46,203 (40.5%) | 7,838 (48.7%) | 0.17 | 2,062 (48.4%) | 0.16 |
| Any comorbidity<sup>c</sup> | 49,201 (43.1%) | 5,844 (36.3%) | 0.14 | 1,898 (44.5%) | 0.03 |
| Number of SARS-CoV-2 tests within 3 months prior to 14 Dec 2020 |  |  |  |  |  |
| 0 | 83,457 (73.2%) | 12,448 (77.4%) | 0.10 | 3,473 (81.5%) | 0.20 |
| 1 | 21,860 (19.2%) | 2,782 (17.3%) | 0.05 | 615 (14.4%) | 0.13 |
| ≥2 | 8,770 (7.7%) | 857 (5.3%) | 0.10 | 173 (4.1%) | 0.15 |
| Receipt of 2019-2020 and/or 2020-2021 influenza vaccination | 40,597 (35.6%) | 4,012 (24.9%) | 0.23 | 1,053 (24.7%) | 0.24 |
| Public health unit region<sup>d</sup> |  |  |  |  |  |
| Central East | 9,139 (8.0%) | 831 (5.2%) | 0.11 | 373 (8.8%) | 0.03 |
| Central West | 21,191 (18.6%) | 3,899 (24.2%) | 0.14 | 864 (20.3%) | 0.04 |
| Durham | 7,690 (6.7%) | 1,050 (6.5%) | 0.01 | 191 (4.5%) | 0.10 |
| Eastern | 7,010 (6.1%) | 865 (5.4%) | 0.03 | 256 (6.0%) | 0.01 |
| North | 7,518 (6.6%) | 265 (1.6%) | 0.25 | 313 (7.3%) | 0.03 |
| Ottawa | 5,716 (5.0%) | 979 (6.1%) | 0.05 | 99 (2.3%) | 0.14 |
| Peel | 12,592 (11.0%) | 2,547 (15.8%) | 0.14 | 526 (12.3%) | 0.04 |
| South West | 12,888 (11.3%) | 998 (6.2%) | 0.18 | 814 (19.1%) | 0.22 |
| Toronto | 21,732 (19.0%) | 3,102 (19.3%) | 0.01 | 542 (12.7%) | 0.17 |
| York | 8,148 (7.1%) | 1,476 (9.2%) | 0.07 | 266 (6.2%) | 0.04 |
| Household income quintile<sup>d, e</sup> |  |  |  |  |  |
| 1 (lowest) | 17,605 (15.4%) | 1,908 (11.9%) | 0.10 | 786 (18.4%) | 0.08 |
| 2 | 20,783 (18.2%) | 2,532 (15.7%) | 0.07 | 784 (18.4%) | 0 |
| 3 | 22,516 (19.7%) | 3,085 (19.2%) | 0.01 | 840 (19.7%) | 0 |
| 4 | 24,861 (21.8%) | 3,773 (23.5%) | 0.04 | 906 (21.3%) | 0.01 |
| 5 (highest) | 27,795 (24.4%) | 4,707 (29.3%) | 0.11 | 925 (21.7%) | 0.06 |
| Essential workers quintile<sup>d, f</sup> |  |  |  |  |  |
| 1 (0%–32.5%) | 26,896 (23.6%) | 4,539 (28.2%) | 0.11 | 648 (15.2%) | 0.21 |
| 2 (32.5%–42.3%) | 28,043 (24.6%) | 4,487 (27.9%) | 0.08 | 973 (22.8%) | 0.04 |
| 3 (42.3%–49.8%) | 22,737 (19.9%) | 3,057 (19.0%) | 0.02 | 903 (21.2%) | 0.03 |
| 4 (50.0%–57.5%) | 19,614 (17.2%) | 2,272 (14.1%) | 0.08 | 834 (19.6%) | 0.06 |
| 5 (57.5%–100%) | 16,034 (14.1%) | 1,621 (10.1%) | 0.12 | 866 (20.3%) | 0.17 |
| Persons per dwelling quintile<sup>d, g</sup> |  |  |  |  |  |
| 1 (0–2.1) | 21,183 (18.6%) | 2,523 (15.7%) | 0.08 | 711 (16.7%) | 0.05 |
| 2 (2.2–2.4) | 19,168 (16.8%) | 1,934 (12.0%) | 0.14 | 738 (17.3%) | 0.01 |
| 3 (2.5–2.6) | 15,197 (13.3%) | 1,919 (11.9%) | 0.04 | 577 (13.5%) | 0.01 |
| 4 (2.7–3.0) | 27,853 (24.4%) | 4,143 (25.8%) | 0.03 | 1,026 (24.1%) | 0.01 |
| 5 (3.1–5.7) | 29,879 (26.2%) | 5,443 (33.8%) | 0.17 | 1,169 (27.4%) | 0.03 |
| Self-identified visible minority quintile<sup>d, h</sup> |  |  |  |  |  |
| 1 (0.0%–2.2%) | 16,930 (14.8%) | 1,439 (8.9%) | 0.18 | 779 (18.3%) | 0.09 |
| 2 (2.2%–7.5%) | 20,294 (17.8%) | 2,235 (13.9%) | 0.11 | 882 (20.7%) | 0.07 |
| 3 (7.5%–18.7%) | 22,577 (19.8%) | 3,408 (21.2%) | 0.03 | 841 (19.7%) | 0 |
| 4 (18.7%–43.5%) | 26,063 (22.8%) | 4,250 (26.4%) | 0.08 | 795 (18.7%) | 0.10 |
| 5 (43.5%–100%) | 27,463 (24.1%) | 4,644 (28.9%) | 0.11 | 928 (21.8%) | 0.05 |
| **Week of test** |  |  |  |  |  |
| 6 Dec 2021 to 12 Dec 2021 | 31,103 (27.3%) | 729 (4.5%) | 0.65 | 1,444 (33.9%) | 0.14 |
| 13 Dec 2021 to 19 Dec 2021 | 41,090 (36.0%) | 5,049 (31.4%) | 0.10 | 1,830 (42.9%) | 0.14 |
| 20 Dec 2021 to 26 Dec 2021 | 41,894 (36.7%) | 10,309 (64.1%) | 0.57 | 987 (23.2%) | 0.30 |
| Prior positive SARS-CoV-2 test | 4,257 (3.7%) | 117 (0.7%) | 0.20 | 10 (0.2%) | 0.25 |
| **COVID-19 vaccine characteristics** |  |  |  |  |  |
| Unvaccinated | 4,681 (4.1%) | 790 (4.9%) | 0.04 | 1,251 (29.4%) | 0.72 |
| Received 2-dose primary series only (with<br>at least 1 mRNA vaccine) | 91,305 (80.0%) | 13,813 (85.9%) | 0.16 | 2,845 (66.8%) | 0.30 |
| Received BNT162b2 for third dose | 14,782 (13.0%) | 1,225 (7.6%) | 0.18 | 134 (3.1%) | 0.37 |
| Received mRNA-1273 for third dose | 3,319 (2.9%) | 259 (1.6%) | 0.09 | 31 (0.7%) | 0.16 |
| **Time since second dose** |  |  |  |  |  |
| 7-59 days | 2,254 (2.0%) | 231 (1.4%) | 0.04 | 61 (1.4%) | 0.04 |
| 60-119 days | 6,769 (5.9%) | 1,003 (6.2%) | 0.01 | 185 (4.3%) | 0.07 |
| 120-179 days | 60,722 (53.2%) | 8,543 (53.1%) | 0 | 1,855 (43.5%) | 0.19 |
| 180-239 days | 19,841 (17.4%) | 3,817 (23.7%) | 0.16 | 668 (15.7%) | 0.05 |
| ≥240 days | 1,719 (1.5%) | 219 (1.4%) | 0.01 | 76 (1.8%) | 0.02 |
| **Time since third dose** |  |  |  |  |  |
| No third dose (i.e., only 2 doses) | 91,305 (80.0%) | 13,813 (85.9%) | 0.16 | 2,845 (66.8%) | 0.30 |
| 0-6 days | 5,963 (5.2%) | 804 (5.0%) | 0.01 | 74 (1.7%) | 0.19 |
| 7-59 days | 11,283 (9.9%) | 638 (4.0%) | 0.23 | 74 (1.7%) | 0.35 |
| ≥60 days | 855 (0.7%) | 42 (0.3%) | 0.07 | 17 (0.4%) | 0.05 |
| **Interval between first and second doses** |  |  |  |  |  |
| 15-34 days | 15,474 (13.6%) | 2,273 (14.1%) | 0.02 | 425 (10.0%) | 0.11 |
| 35-55 days | 37,972 (33.3%) | 6,154 (38.3%) | 0.10 | 988 (23.2%) | 0.23 |
| ≥56 days | 55,960 (49.1%) | 6,870 (42.7%) | 0.13 | 1,597 (37.5%) | 0.24 |
| **Interval between second and third doses** |  |  |  |  |  |
| No third dose (i.e., only 2 doses) | 91,305 (80.0%) | 13,813 (85.9%) | 0.16 | 2,845 (66.8%) | 0.30 |
| ≤111 days | 232 (0.2%) | 18 (0.1%) | 0.02 | 8 (0.2%) | 0 |
| 112-167 days | 1,617 (1.4%) | 122 (0.8%) | 0.06 | 26 (0.6%) | 0.08 |
| ≥168 days | 16,252 (14.2%) | 1,344 (8.4%) | 0.19 | 131 (3.1%) | 0.41 |

<sup>a</sup>Proportion reported, unless stated otherwise.
<sup>b</sup>SD=standardized difference. Standardized differences of >0.10 are considered clinically relevant. Comparison of Omicron-positive cases with SARS-CoV-2-negative controls, and Delta-positive cases with SARS-CoV-2-negative controls.
<sup>c</sup>Comorbidities include chronic respiratory diseases, chronic heart diseases, hypertension, diabetes, immunocompromising conditions due to underlying diseases or therapy, autoimmune diseases, chronic kidney disease, advanced liver disease, dementia/frailty and history of stroke or transient ischemic attack.
<sup>d</sup>The sum of counts does not equal the column total because of individuals with missing information (<1.0%) for this characteristic.
<sup>e</sup>Household income quintile has variable cut-off values in each city/Census area to account for cost of living. A dissemination area (DA) being in quintile 1 means it is among the lowest 20% of DAs in its city by income.
<sup>f</sup>Percentage of people in the area working in the following occupations: sales and service occupations; trades, transport and equipment operators and related occupations; natural resources, agriculture, and related production occupations; and occupations in manufacturing and utilities. Census counts for people are randomly rounded up or down to the nearest number divisible by 5, which causes some minor imprecision.
<sup>g</sup>Range of persons per dwelling.
<sup>h</sup>Percentage of people in the area who self-identified as a visible minority. Census counts for people are randomly rounded up or down to the nearest number divisible by 5, which causes some minor imprecision.

In contrast, Delta cases were more similar to controls than were Omicron cases in some respects (e.g., age, comorbidities) but were more different in others, such as being more likely to have occurred earlier in the study period, and more likely to be unvaccinated (29.4% vs. 4.1%).

In the Omicron VE analyses, vaccinated (i.e., 2 or 3 dose) subjects were more likely to be older, female, influenza vaccine recipients, and residents of neighbourhoods with higher income and fewer essential workers than unvaccinated subjects (Table 2). Recipients of a third dose were more likely to have comorbidities and to have been tested in the last week of the study period. We observed similar patterns for subjects in the Delta VE analyses (Supplementary Table 1).

**Table 2.**
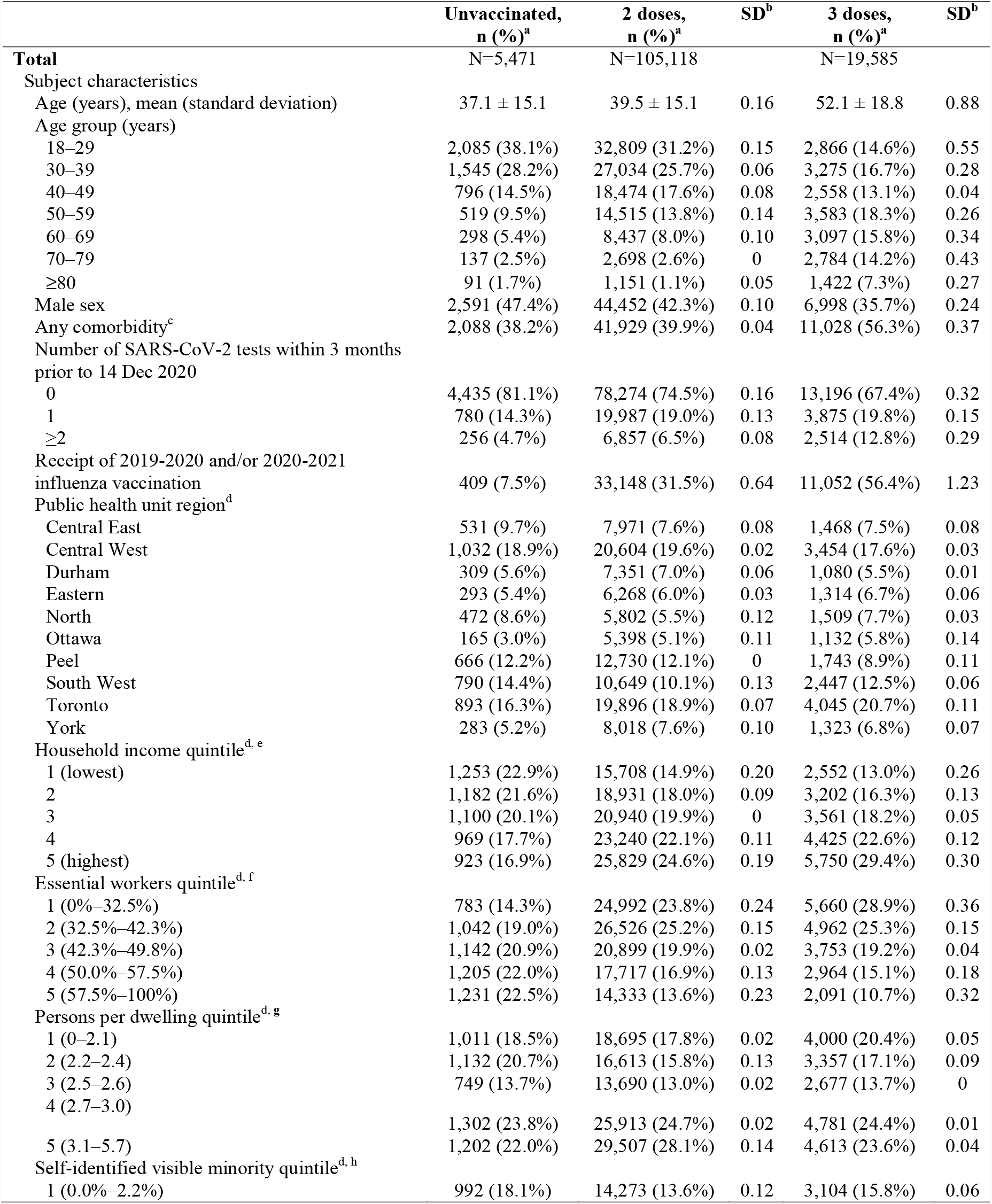
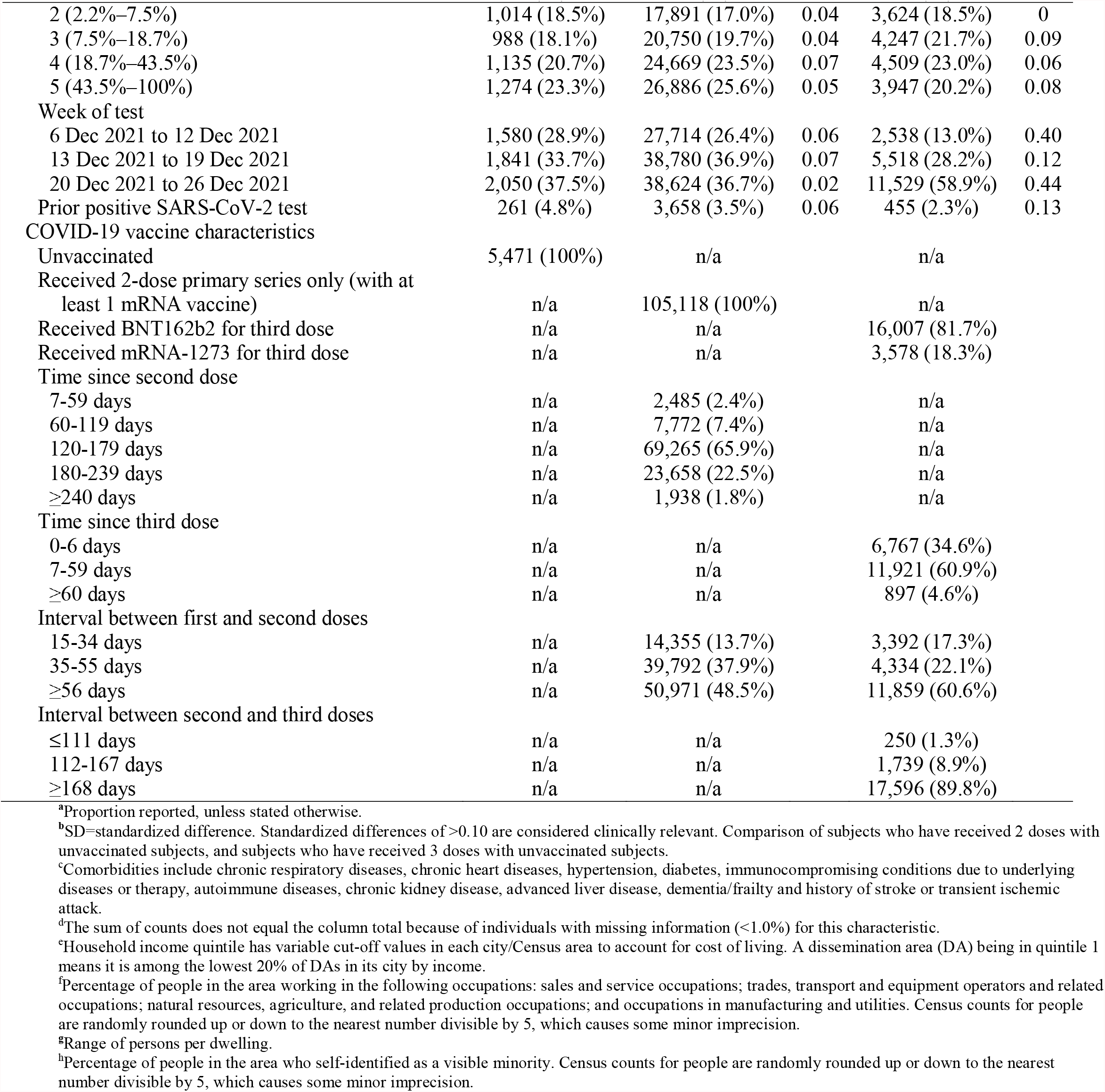
Descriptive characteristics of subjects tested for SARS-CoV-2 with COVID-19-relevant symptoms during the period December 6 to 26, 2021, comparing vaccinated and unvaccinated subjects, Omicron cases and SARS-CoV-2-negative controls only

After 2 doses of COVID-19 vaccines (with at least 1 mRNA vaccine), VE against symptomatic Delta infection declined steadily over time from 89% (95%CI, 86-92%) 7-59 days after a second dose to 80% (95%CI, 74-84%) after ≥240 days, but increased to 97% (95%CI, 96-98%) ≥7 days after a third dose (Table 3; Figure 1A). VE against symptomatic Omicron infection was lower relative to Delta for the entire period and waned more rapidly, from 36% (95%CI, 24-45%) 7-59 days after a second dose to 1% (95%CI, –8% to 10%) 180-239 days after a second dose. VE against symptomatic Omicron infection was 61% (95%CI, 56-65%) ≥7 days after a third dose and was similar regardless of the mRNA product administered.

**Table 3.** Estimates of vaccine effectiveness* against symptomatic infection or severe outcomes (hospitalization or death) caused by Omicron or Delta during the period December 6 to 26, 2021, by time since latest dose

| Outcome | Doses | Vaccine products | Days since latest dose | SARS-CoV-2 negative, n | Omicron, n | Vaccine effectiveness against Omicron (95% CI) | Delta, n | Vaccine effectiveness against Delta (95% CI) |
| --- | --- | --- | --- | --- | --- | --- | --- | --- |
| Symptomatic infection | First 2 doses | ≥1 mRNA vaccine | 7-59 | 2254 | 231 | 36 (24, 45) | 61 | 89 (86, 92) |
|  |  |  | 60-119 | 6769 | 1003 | 12 (3, 21) | 185 | 89 (87, 90) |
|  |  |  | 120-179 | 60722 | 8543 | 15 (8, 22) | 1855 | 88 (87, 89) |
|  |  |  | 180-239 | 19841 | 3817 | 1 (-8, 10) | 668 | 85 (83, 87) |
|  |  |  | ≥240 | 1719 | 219 | 2 (-17, 17) | 76 | 80 (74, 84) |
|  | Third dose | Any mRNA vaccine | 0-6 | 5963 | 804 | 36 (29, 43) | 74 | 94 (93, 95) |
|  |  |  | ≥7 | 12138 | 680 | 61 (56, 65) | 91 | 97 (96, 98) |
|  |  | BNT162b2 | 0-6 | 4509 | 625 | 34 (26, 41) | 60 | 94 (92, 95) |
|  |  |  | ≥7 | 10273 | 600 | 60 (55, 65) | 74 | 97 (96, 98) |
|  |  | mRNA-1273 | 0-6 | 1454 | 179 | 43 (32, 52) | 14 | 95 (92, 97) |
|  |  |  | ≥7 | 1865 | 80 | 65 (55, 72) | 17 | 97 (95, 98) |
| Severe outcomes | First 2 doses | ≥1 mRNA vaccine | 7-59 | 2254 | ≤5 | 55 (-106, 90) | ≤5 | 94 (84, 98) |
|  |  |  | 60-119 | 6769 | 6 | 37 (-71, 77) | ≤5 | 98 (94, 99) |
|  |  |  | 120-179 | 60722 | 31 | 75 (51, 87) | 40 | 99 (98, 99) |
|  |  |  | 180-239 | 19841 | 18 | 82 (62, 91) | 32 | 97 (96, 98) |
|  |  |  | ≥240 | 1719 | ≤5 | 86 (-12, 98) | ≤5 | 95 (85, 99) |
|  | Third dose | Any mRNA vaccine | 0-6 | 5963 | ≤5 | 91 (71, 97) | 6 | 99 (97, 99) |
|  |  |  | ≥7 | 12138 | 11 | 95 (87, 98) | 16 | 99 (98, 99) |
|  |  | BNT162b2 | 0-6 | 4509 | ≤5 | 88 (62, 96) | 6 | 98 (96, 99) |
|  |  |  | ≥7 | 10273 | 8 | 95 (87, 98) | 15 | 99 (98, 99) |
|  |  | mRNA-1273 | 0-6 | 1454 | 0 | ¶ | 0 | ¶ |
|  |  |  | ≥7 | 1865 | ≤5 | 93 (74, 98) | ≤5 | 100 (98, 100) |

\*Vaccine effectiveness estimates adjusted for: age (in 10-year age bands), sex, public health unit region of residence, number of SARS-CoV-2 PCR tests during the 3 months prior to December 14, 2020, past SARS-CoV-2 infection >90 days prior to index date, comorbidities, influenza vaccination status during the 2019/2020 and/or 2020/2021 influenza seasons, and neighbourhood-level information on median household income, proportion of the working population employed as non-health essential workers, mean number of persons per dwelling, and proportion of the population who self-identify as a visible minority.
¶Vaccine effectiveness estimated as 100% based on zero vaccinated test-positive cases.

**Figure 1.**
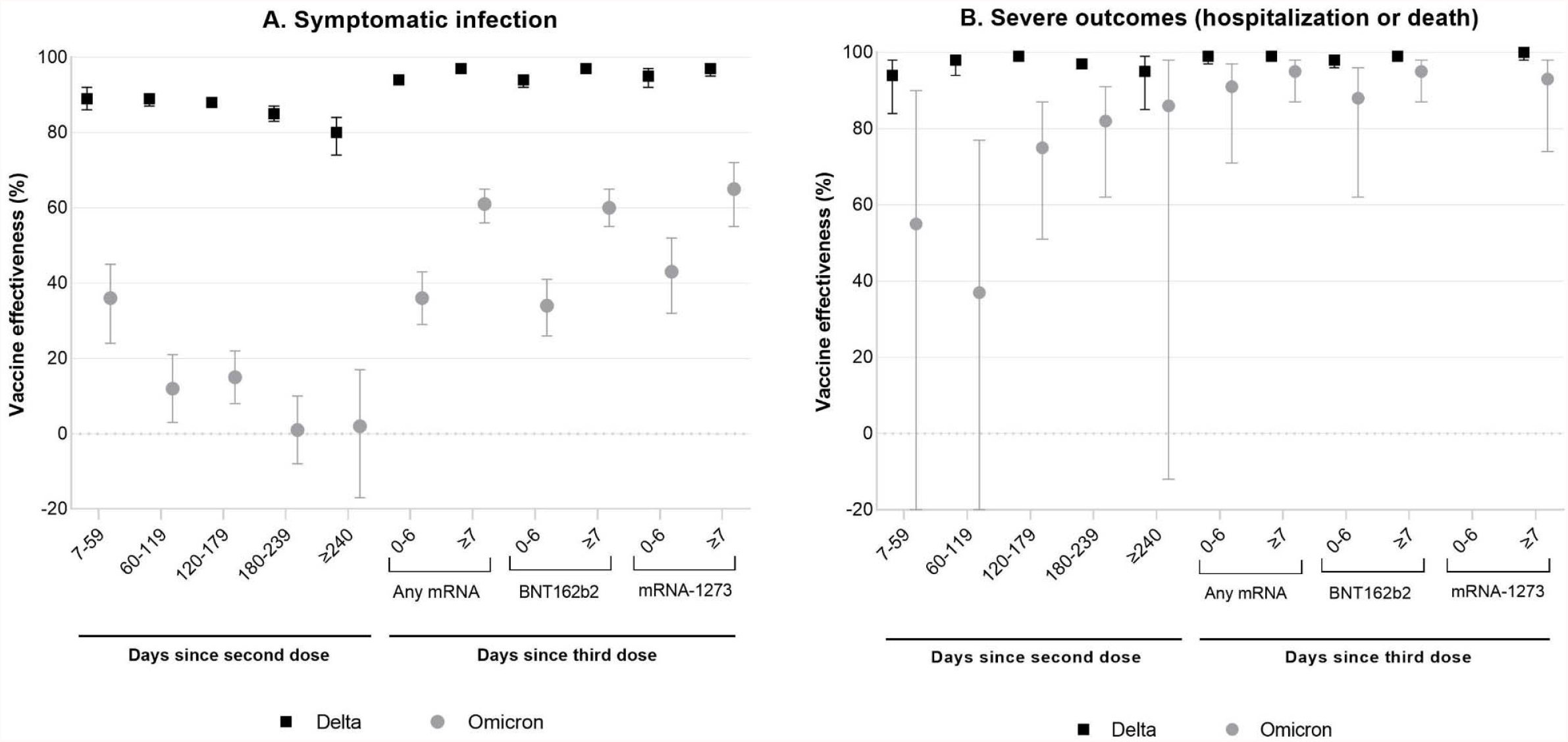
Estimates of vaccine effectiveness against A) symptomatic infection and B) severe outcomes (hospitalization or death) caused by Omicron or Delta during the period December 6 to 26, 2021, by time since latest dose Vaccine effectiveness (VE) for mRNA-1273 0-6 days after the third dose was estimated as 100% based on zero vaccinated test-positive hospitalized cases and was therefore not presented in panel B. The lower 95% confidence limit for Omicron VE against severe outcomes 7-59 days after a second dose was -106 and 60-119 days after a second dose was -71.

VE was higher against severe outcomes when compared to symptomatic infection and did not exhibit the same degree of waning (Figure 1B). VE was also higher against severe outcomes caused by Delta than Omicron in the time following a second dose, but confidence intervals were very wide for some Omicron estimates. VE against severe outcomes ≥7 days following a third dose were very similar for Delta and Omicron (99% [95%CI, 98-99%] and 95% [95%CI, 87-98%], respectively).

In the sensitivity analysis examining cumulative changes in VE over calendar time, point estimates generally rose and confidence intervals narrowed as more data accumulated (Figure 2). There were some changes in the characteristics of Omicron cases over time, including an increase in mean age (from 32.0 years in the first 3 weeks after Omicron emergence to 36.3 years in the final week of our study), and a decrease in the proportion of subjects who resided in the highest neighbourhood income quintile (from 35.4% to 27.8%) (Supplementary Table 2). When using a less specific definition for Omicron (Supplementary Table 3), VE estimates against severe outcomes were higher and substantially more stable (Supplementary Table 4).

**Figure 2.**
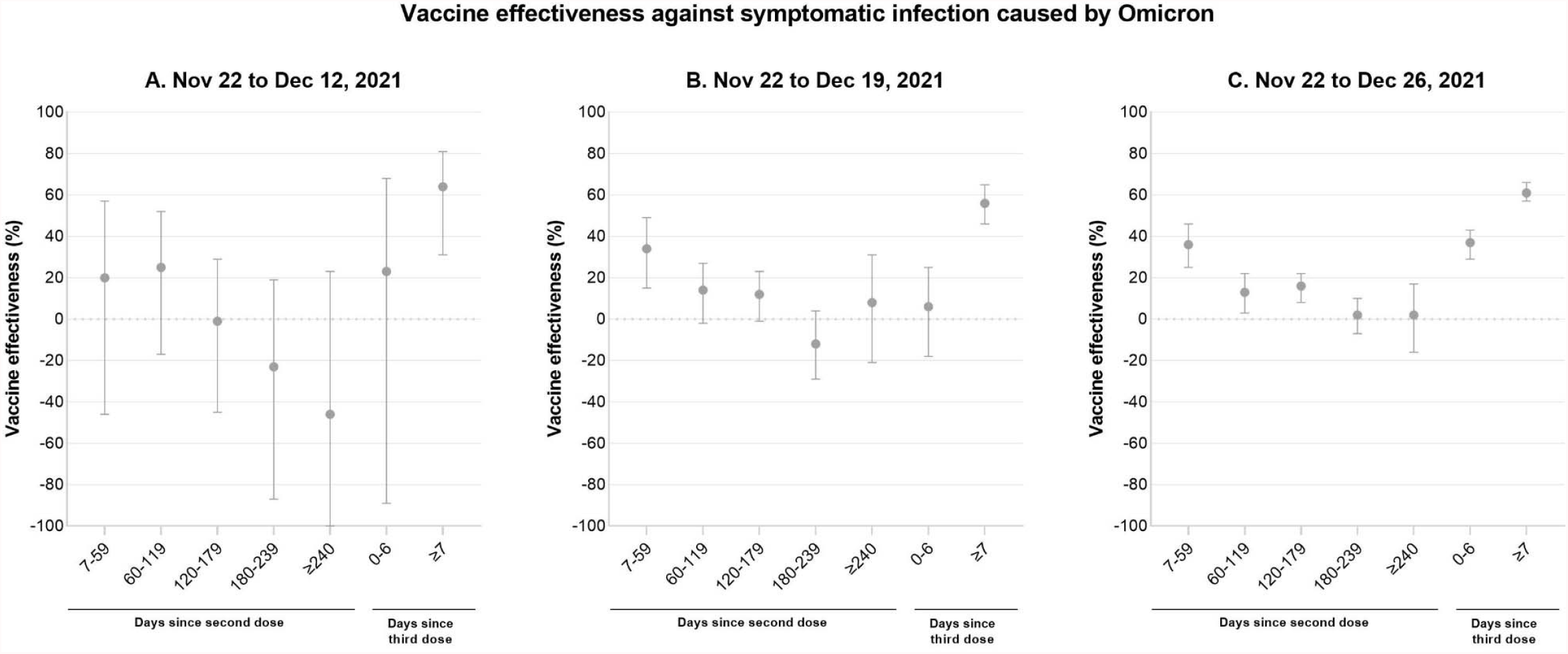
Estimates of vaccine effectiveness (VE) against symptomatic Omicron infection by cumulative time period to demonstrate the impact of adding data from successive weeks to VE estimates (panel A: November 22 to December 21, 2021; panel B: November 22 to December 19, 2021; panel C: November 22 to December 26, 2021) The lower 95% confidence limit for vaccine effectiveness against symptomatic Omicron infection ≥240 days after a second dose was -174

In our supplemental analysis of VE against any infection (irrespective of symptoms and severity), we found very similar results to our earlier estimates but with narrower confidence intervals when using the new study period (Supplementary Figure 1).

## DISCUSSION

In Ontario, between December 6 and 26, 2021, we found the effectiveness of 2 doses of COVID-19 vaccines against symptomatic infection to be substantially lower for Omicron than for Delta; VE against Omicron symptomatic infection was minimal from 60 days after a second dose and there was no significant protection beyond 180 days. However, VE was 61% ≥7 days following a third dose. Against severe outcomes, including hospitalization and death, VE shortly following a third dose was similar for Omicron and Delta (95% [95%CI 87-98%] and 99% [95% CI 98-99%], respectively).

Substantial waning of 2-dose VE against symptomatic infection has been observed in England, with lower VE against Omicron than Delta at each interval following 2 or 3 doses;^11 13^ our results demonstrated a similar pattern, with a marked reduction in 2-dose (with at least one being an mRNA vaccine) and 3-dose effectiveness against symptomatic Omicron infection relative to Delta. In the United Kingdom Health Security Agency (UKHSA) data, VE against symptomatic Omicron infection for 2 doses of BNT162b2 or mRNA-1273 declined to <20% from 20 weeks after a second dose. Following a third dose of mRNA vaccine, VE increased to ∼60-75% during the first 4 weeks, in line with our finding of 61% ≥7 days after a third dose.^11^ While the UK data suggest waning also occurs after a dose, insufficient time has elapsed for enough third dose recipients in Ontario to assess this. In Scotland, a third dose was associated with a two-thirds reduction in the odds of symptomatic Omicron infection relative to those who were ≥25 weeks post second dose in the early Omicron period.^15^

Although prior studies have demonstrated reduced neutralizing antibodies against Omicron relative to other variants following receipt of 2 mRNA vaccine doses^4-8 32 33^ (but with potent neutralization following a third dose^34 35 36^), CD8+ cytotoxic T cells are less impacted by mutations in the Omicron variant and likely continue to provide protection against severe disease.^35 37^ Emerging real-world data demonstrate that protection against severe outcomes is more preserved than against infection in the Omicron era. In South Africa, effectiveness against hospitalization was 93% in the pre-Omicron period and was 70% in the Omicron period.^16^ In England, VE against hospitalization due to Omicron also appears to be better maintained relative to symptomatic infection with Omicron.^10 11 38^ In an analysis of the overall population and for all vaccines (i.e., BNT162b2, mRNA-1273, and ChAdOx1), VE against Omicron-related hospitalization declined with time since a second dose but to a lesser extent than against symptomatic infection.^11^ Following a third dose, VE was restored to 85-95% during the first 3 months. In California, VE against Omicron-related hospitalization was 89% in the first 3 months following 3 doses of BNT162b2.^12^ In another study in the United States (US), 3-dose VE against hospitalization when Omicron was predominant was estimated at 90%.^17^ Our 3-dose estimates against severe outcomes were very similar to these results from England and the US. Although our 2-dose estimates against severe Omicron-related outcomes were challenging to compare given instability and wide confidence intervals, they do suggest less waning against hospitalization than has been observed elsewhere and this finding is aligned with surveillance data in Ontario.^39 40^

Direct comparisons to other jurisdictions are challenging^41^ due to differences in study methodology, outcome definitions (i.e., identification of Omicron and Delta based on laboratory confirmation vs. time-based criteria), intervals following latest dose selected to monitor VE, vaccination policies (i.e., homologous vs. heterologous vaccine schedules, third dose eligibility criteria, product-specific policies [use of mRNA vs. viral vector vaccines, a preferential recommendation in Ontario of BNT162b2 for young adults]),^42 43^ population age structures, public health measures implemented during the study period (e.g., vaccine certificates, mask mandates^44^), testing patterns, and use of antivirals or other therapies. Further, Ontario has experienced a lower cumulative incidence of reported infections and has attained higher vaccine coverage than other countries that have estimated VE against Omicron to date,^45^ and thus has a potentially dissimilar distribution of infection-induced versus vaccine-induced immunity.^45 46^ Despite this, the general trends across the studies are similar and suggest immune evasion by Omicron.^10-12 15 17 18 47^

In our earlier analysis encompassing the first month of Omicron emergence in Ontario, we estimated VE against infection (irrespective of presence of symptoms or severity, due to the data availability at the time) and observed statistically significant negative VE estimates for some time intervals following a second dose.^19^ This was also observed in the UK (estimated against symptomatic infection) for ChAdOx1 recipients and Denmark (estimated against any infection), where analyses were conducted promptly after Omicron emergence.^13 14 48 49^ As more data accumulated in the UK, updated results demonstrated that while protection was still minimal for longer periods after a second dose, the VE estimates were no longer negative.^11 38^ In Ontario, the initial negative VE estimates were likely due to a combination of factors. First, a vaccine certificate system was introduced in Ontario in the fall of 2021, such that only individuals who have received 2 vaccine doses are permitted to travel by air and rail, and to enter restaurants, bars, gyms, and large cultural and sporting events. Introduction of Omicron into Ontario by vaccinated travellers and the initial spread via household and social contacts (who are also more likely to be vaccinated) likely resulted in increased risk of exposure to Omicron among vaccinated individuals before gradually diffusing into the broader population, including unvaccinated individuals. This hypothesis is supported by earlier Omicron cases being younger, residing in higher-income neighbourhoods, and being more highly vaccinated than the test-negative controls during the same period. In subsequent weeks, cases became more similar to test-negative controls in terms of age, presence of comorbidities, and sociodemographic measures; the proportion unvaccinated also increased over time among cases, eventually becoming higher than among test-negative controls. Second, use of infection rather than symptomatic infection as the outcome, as in the initial analysis,^19^ may also, introduce bias in VE estimates because indications for testing (e.g., contact with a case) may be differential by vaccination status. If asymptomatic vaccinated individuals are more likely to be tested (e.g., healthcare workers, individuals exposed to cases), vaccinated Omicron cases would be more likely to be identified compared to unvaccinated Omicron cases, leading to an underestimate of VE.^50^ Restricting the analysis to symptomatic individuals, as presented here, should mitigate this potential source of bias.

Our study has several limitations. First, we were unable to differentiate individuals who received a third dose as part of an extended primary series (i.e., severely or moderately immunocompromised individuals) as well as higher-risk individuals who were eligible for a third dose earlier (e.g., residents of retirement homes). As third dose eligibility only expanded to all adults in mid-December, a proportion of subjects in this study who had a third dose may reflect these highly vulnerable populations; thus, our 3-dose VE estimates may be lower than what would be expected for the general population. Third dose eligibility and timing of primary series completion may result in differing characteristics of individuals by interval since last dose. Second, not all specimens were screened for SGTF due to the rapid rise in case volumes and eligibility criteria for screening, and WGS results for cases identified during our study period may not have been available at the time of analysis due to processing delays. In a sensitivity analysis for severe outcomes we obtained a higher VE estimate when we classified all cases after a certain date as Omicron. However, this less specific outcome may over-estimate VE if there is differential misclassification of the variant based on vaccination status (i.e., if Delta continues to circulate at a higher level in unvaccinated individuals, this would over-estimate VE). Third, changes in testing patterns, including increased use of rapid antigen tests (which are not captured in our data) and decreased PCR testing availability, may have impacted our estimates, but the direction of any resulting bias is uncertain. However, if vaccinated individuals are more likely to be tested and therefore captured in our data than unvaccinated individuals, then VE will be underestimated.^50^ Fourth, symptom information is not available for all laboratories submitting to Ontario’s centralized system; as such, symptomatic individuals who were tested but without this information recorded would have been excluded in our study.^23^ Fifth, despite ongoing high case counts, we were unable to extend our time period past the end of December due to restricted test eligibility and access, reduced SGTF screening and the likelihood of misclassification due to the potential rise of the BA.2 sub-lineage of Omicron, as noted elsewhere,^38 51^ but so far has been minimal in Ontario. Sixth, we were unable to estimate VE in the extended period of time following a third dose. Last, there may be residual confounding that was not accounted for in our analysis. This includes an inability to control for previous undocumented infections, which may be differential by vaccination status, and confounding due to behavioural patterns.

Our results demonstrate the importance of third doses to bolster protection against both infections and severe outcomes due to Omicron, although the duration of this protection is uncertain. They also suggest the continued need for public health measures, such as masking, physical distancing, and ventilation, to prevent infection and transmission. Further, these findings have potentially important implications for proof of vaccination requirements, in that 3 doses are necessary to reduce transmission in high-risk settings although they do not confer as much protection as observed against Delta. Our work adds to a rapidly evolving body of evidence that suggests that vaccine-induced protection depends on a variety of factors such as type of vaccine received, recipient age, time since latest dose, and circulating variant.

## Conclusions

Our results suggest that protection from 2 doses of COVID-19 vaccines against symptomatic Omicron infection declines with time since a second dose, with no protection beyond 180 days, and is substantially lower than against Delta infection; a third dose of mRNA vaccine affords moderate protection against symptomatic Omicron infection in the immediate term. In contrast, protection against Omicron-related severe disease appears to be high with 2 doses and is further increased and is similar to Delta with a third dose. Additional tools beyond the currently available vaccines, such as ongoing public health measures, antivirals or other therapies, and new formulations of COVID-19 vaccines, are likely needed to mitigate the impact of Omicron and potential future variants.

## Data Availability

The dataset from this study is held securely in coded form at ICES. While legal data sharing agreements between ICES and data providers (e.g., healthcare organizations and government) prohibit ICES from making the dataset publicly available, access may be granted to those who meet pre-specified criteria for confidential access, available at www.ices.on.ca/DAS.

## Ethics approval

ICES is a prescribed entity under Ontario’s Personal Health Information Protection Act (PHIPA). Section 45 of PHIPA authorizes ICES to collect personal health information, without consent, for the purpose of analysis or compiling statistical information with respect to the management of, evaluation or monitoring of, the allocation of resources to or planning for all or part of the health system. Projects that use data collected by ICES under section 45 of PHIPA, and use no other data, are exempt from REB review. The use of the data in this project is authorized under section 45 and approved by ICES’ Privacy and Legal Office.

## Code availability

The full dataset creation plan and underlying analytic code are available from the authors upon request, understanding that the computer programs may rely upon coding templates or macros that are unique to ICES and are therefore either inaccessible or may require modification.

## Acknowledgments

We would like to acknowledge Public Health Ontario for access to vaccination data from COVaxON, case-level data from CCM and COVID-19 laboratory data, as well as assistance with data interpretation. We also thank the staff of Ontario’s public health units who are responsible for COVID-19 case and contact management and data collection within CCM. We thank IQVIA Solutions Canada Inc. for use of their Drug Information Database. The authors are grateful to the Ontario residents without whom this research would be impossible.

## Author contributions

S.A.B, H.C., and J.C.K. designed the study. H.C. obtained the data and conducted all analyses (data set and variable creation and statistical modelling). S.A.B. and J.C.K. drafted the manuscript. All authors contributed to the analysis plan, interpreted the results, critically reviewed and edited the manuscript, approved the final version, and agreed to be accountable for all aspects of the work.

## Funding and disclaimers

This work was supported by the Canadian Immunization Research Network (CIRN) through a grant from the Public Health Agency of Canada and the Canadian Institutes of Health Research (CNF 151944). This project was also supported by funding from the Public Health Agency of Canada, through the Vaccine Surveillance Reference Group and the COVID-19 Immunity Task Force. This study was also supported by ICES, which is funded by an annual grant from the Ontario Ministry of Health (MOH). J.C.K. is supported by Clinician-Scientist Award from the University of Toronto Department of Family and Community Medicine. P.C.A. is supported by a Mid-Career Investigator Award from the Heart and Stroke Foundation.

This work was supported by Public Health Ontario. This study was also supported by ICES, which is funded by an annual grant from the Ontario Ministry of Health (MOH) and the Ministry of Long-Term Care (MLTC). This study was supported by the Ontario Health Data Platform (OHDP), a Province of Ontario initiative to support Ontario’s ongoing response to COVID-19 and its related impacts. The study sponsors did not participate in the design and conduct of the study; collection, management, analysis and interpretation of the data; preparation, review or approval of the manuscript; or the decision to submit the manuscript for publication. Parts of this material are based on data and/or information compiled and provided by the Canadian Institute for Health Information (CIHI) and by Cancer Care Ontario (CCO). However, the analyses, conclusions, opinions and statements expressed herein are solely those of the authors, and do not reflect those of the funding or data sources; no endorsement by ICES, MOH, MLTC, OHDP, its partners, the Province of Ontario, CIHI or CCO is intended or should be inferred.

