## Supplementary materials for "Effectiveness of COVID-19 vaccines against Omicron or Delta symptomatic infection and severe outcomes"

**Supplementary Table 1.** Descriptive characteristics of subjects tested for SARS-CoV-2 with COVID-19-relevant symptoms during the period December 6 to 26, 2021, comparing vaccinated and unvaccinated subjects, Delta cases and SARS-CoV-2-negative controls only

|  | **Unvaccinated, n (%)^a^** | **2 doses,**  **n (%)^a^** | **SD^b^** | **3 doses,**  **n (%)^a^** | **SD^b^** |
| --- | --- | --- | --- | --- | --- |
| **Total** | N=5,932 | N=94,150 |  | N=18,266 |  |
| Subject characteristics |  |  |  |  |  |
| Age (years), mean (standard deviation) | 38.9 ± 15.8 | 40.2 ± 15.3 | 0.09 | 52.6 ± 18.8 | 0.79 |
| Age group (years) |  |  |  |  |  |
| 18–29 | 2,016 (34.0%) | 27,376 (29.1%) | 0.11 | 2,515 (13.8%) | 0.49 |
| 30–39 | 1,637 (27.6%) | 24,615 (26.1%) | 0.03 | 3,076 (16.8%) | 0.26 |
| 40–49 | 902 (15.2%) | 16,612 (17.6%) | 0.07 | 2,344 (12.8%) | 0.07 |
| 50–59 | 645 (10.9%) | 13,457 (14.3%) | 0.10 | 3,272 (17.9%) | 0.20 |
| 60–69 | 423 (7.1%) | 8,188 (8.7%) | 0.06 | 2,948 (16.1%) | 0.28 |
| 70–79 | 199 (3.4%) | 2,738 (2.9%) | 0.03 | 2,718 (14.9%) | 0.41 |
| ≥80 | 110 (1.9%) | 1,164 (1.2%) | 0.05 | 1,393 (7.6%) | 0.27 |
| Male sex | 2,825 (47.6%) | 39,011 (41.4%) | 0.12 | 6,429 (35.2%) | 0.25 |
| Any comorbidity^c^ | 2,396 (40.4%) | 38,250 (40.6%) | 0 | 10,453 (57.2%) | 0.34 |
| Number of SARS-CoV-2 tests within 3 months prior to 14 Dec 2020 |  |  |  |  |  |
| 0 | 4,847 (81.7%) | 69,875 (74.2%) | 0.18 | 12,208 (66.8%) | 0.35 |
| 1 | 825 (13.9%) | 17,984 (19.1%) | 0.14 | 3,666 (20.1%) | 0.16 |
| ≥2 | 260 (4.4%) | 6,291 (6.7%) | 0.10 | 2,392 (13.1%) | 0.31 |
| Receipt of 2019-2020 and/or 2020-2021 influenza vaccination | 455 (7.7%) | 30,775 (32.7%) | 0.66 | 10,420 (57.0%) | 1.24 |
| Public health unit region^d^ |  |  |  |  |  |
| Central East | 616 (10.4%) | 7,487 (8.0%) | 0.08 | 1,409 (7.7%) | 0.09 |
| Central West | 1,123 (18.9%) | 17,822 (18.9%) | 0 | 3,110 (17.0%) | 0.05 |
| Durham | 299 (5.0%) | 6,573 (7.0%) | 0.08 | 1,009 (5.5%) | 0.02 |
| Eastern | 325 (5.5%) | 5,707 (6.1%) | 0.02 | 1,234 (6.8%) | 0.05 |
| North | 568 (9.6%) | 5,768 (6.1%) | 0.13 | 1,495 (8.2%) | 0.05 |
| Ottawa | 153 (2.6%) | 4,635 (4.9%) | 0.12 | 1,027 (5.6%) | 0.15 |
| Peel | 649 (10.9%) | 10,922 (11.6%) | 0.02 | 1,547 (8.5%) | 0.08 |
| South West | 1,035 (17.4%) | 10,304 (10.9%) | 0.19 | 2,363 (12.9%) | 0.13 |
| Toronto | 858 (14.5%) | 17,617 (18.7%) | 0.11 | 3,799 (20.8%) | 0.17 |
| York | 267 (4.5%) | 6,941 (7.4%) | 0.12 | 1,206 (6.6%) | 0.09 |
| Household income quintile^d, e^ |  |  |  |  |  |
| 1 (lowest) | 1,418 (23.9%) | 14,532 (15.4%) | 0.21 | 2,441 (13.4%) | 0.27 |
| 2 | 1,295 (21.8%) | 17,245 (18.3%) | 0.09 | 3,027 (16.6%) | 0.13 |
| 3 | 1,179 (19.9%) | 18,816 (20.0%) | 0 | 3,361 (18.4%) | 0.04 |
| 4 | 1,071 (18.1%) | 20,620 (21.9%) | 0.10 | 4,076 (22.3%) | 0.11 |
| 5 (highest) | 922 (15.5%) | 22,528 (23.9%) | 0.21 | 5,270 (28.9%) | 0.32 |
| Essential workers quintile^d, f^ |  |  |  |  |  |
| 1 (0%–32.5%) |  |  |  |  |  |
| 2 (32.5%–42.3%) | 732 (12.3%) | 21,601 (22.9%) | 0.28 | 5,211 (28.5%) | 0.41 |
| 3 (42.3%–49.8%) | 1,042 (17.6%) | 23,401 (24.9%) | 0.18 | 4,573 (25.0%) | 0.18 |
| 4 (50.0%–57.5%) | 1,245 (21.0%) | 18,893 (20.1%) | 0.02 | 3,502 (19.2%) | 0.05 |
| 5 (57.5%–100%) | 1,365 (23.0%) | 16,254 (17.3%) | 0.14 | 2,829 (15.5%) | 0.19 |
| Persons per dwelling quintile^d,^ **^g^** |  |  |  |  |  |
| 1 (0–2.1) | 1,120 (18.9%) | 17,004 (18.1%) | 0.02 | 3,770 (20.6%) | 0.04 |
| 2 (2.2–2.4) | 1,261 (21.3%) | 15,453 (16.4%) | 0.12 | 3,192 (17.5%) | 0.10 |
| 3 (2.5–2.6) | 833 (14.0%) | 12,429 (13.2%) | 0.02 | 2,512 (13.8%) | 0.01 |
| 4 (2.7–3.0) | 1,424 (24.0%) | 23,016 (24.4%) | 0.01 | 4,439 (24.3%) | 0.01 |
| 5 (3.1–5.7) | 1,210 (20.4%) | 25,632 (27.2%) | 0.16 | 4,206 (23.0%) | 0.06 |
| Self-identified visible minority quintile^d, h^ |  |  |  |  |  |
| 1 (0.0%–2.2%) | 1,225 (20.7%) | 13,526 (14.4%) | 0.17 | 2,958 (16.2%) | 0.12 |
| 2 (2.2%–7.5%) | 1,186 (20.0%) | 16,549 (17.6%) | 0.06 | 3,441 (18.8%) | 0.03 |
| 3 (7.5%–18.7%) | 1,052 (17.7%) | 18,445 (19.6%) | 0.05 | 3,921 (21.5%) | 0.09 |
| 4 (18.7%–43.5%) | 1,139 (19.2%) | 21,562 (22.9%) | 0.09 | 4,157 (22.8%) | 0.09 |
| 5 (43.5%–100%) | 1,251 (21.1%) | 23,495 (25.0%) | 0.09 | 3,645 (20.0%) | 0.03 |
| Week of test |  |  |  |  |  |
| 6 Dec 2021 to 12 Dec 2021 | 1,999 (33.7%) | 27,997 (29.7%) | 0.09 | 2,551 (14.0%) | 0.48 |
| 13 Dec 2021 to 19 Dec 2021 | 2,152 (36.3%) | 35,471 (37.7%) | 0.03 | 5,297 (29.0%) | 0.16 |
| 20 Dec 2021 to 26 Dec 2021 | 1,781 (30.0%) | 30,682 (32.6%) | 0.06 | 10,418 (57.0%) | 0.57 |
| Prior positive SARS-CoV-2 test | 253 (4.3%) | 3,568 (3.8%) | 0.02 | 446 (2.4%) | 0.10 |
| COVID-19 vaccine characteristics |  |  |  |  |  |
| Unvaccinated | 5,932 (100%) | n/a |  | n/a |  |
| Received 2-dose primary series only (with at least 1 mRNA vaccine) | n/a | 94,150 (100%) |  | n/a |  |
| Received BNT162b2 for third dose | n/a | n/a |  | 14,916 (81.7%) |  |
| Received mRNA-1273 for third dose | n/a | n/a |  | 3,350 (18.3%) |  |
| Time since second dose |  |  |  |  |  |
| 7-59 days | n/a | 2,315 (2.5%) |  | n/a |  |
| 60-119 days | n/a | 6,954 (7.4%) |  | n/a |  |
| 120-179 days | n/a | 62,577 (66.5%) |  | n/a |  |
| 180-239 days | n/a | 20,509 (21.8%) |  | n/a |  |
| ≥240 days | n/a | 1,795 (1.9%) |  | n/a |  |
| Time since third dose |  |  |  |  |  |
| 0-6 days | n/a | n/a |  | 6,037 (33.1%) |  |
| 7-59 days | n/a | n/a |  | 11,357 (62.2%) |  |
| ≥60 days | n/a | n/a |  | 872 (4.8%) |  |
| Interval between first and second doses |  |  |  |  |  |
| 15-34 days | n/a | 12,675 (13.5%) |  | 3,224 (17.7%) |  |
| 35-55 days | n/a | 34,995 (37.2%) |  | 3,965 (21.7%) |  |
| ≥56 days | n/a | 46,480 (49.4%) |  | 11,077 (60.6%) |  |
| Interval between second and third doses |  |  |  |  |  |
| ≤111 days | n/a | n/a |  | 240 (1.3%) |  |
| 112-167 days | n/a | n/a |  | 1,643 (9.0%) |  |
| ≥168 days | n/a | n/a |  | 16,383 (89.7%) |  |

**^a^**Proportion reported, unless stated otherwise.

**^b^**SD=standardized difference. Standardized differences of >0.10 are considered clinically relevant. Comparison of subjects who have received 2 doses with unvaccinated subjects, and subjects who have received 3 doses with unvaccinated subjects.

^c^Comorbidities include chronic respiratory diseases, chronic heart diseases, hypertension, diabetes, immunocompromising conditions due to underlying diseases or therapy, autoimmune diseases, chronic kidney disease, advanced liver disease, dementia/frailty and history of stroke or transient ischemic attack.

^d^The sum of counts does not equal the column total because of individuals with missing information (<1.0%) for this characteristic.

^e^Household income quintile has variable cut-off values in each city/Census area to account for cost of living. A dissemination area (DA) being in quintile 1 means it is among the lowest 20% of DAs in its city by income.

^f^Percentage of people in the area working in the following occupations: sales and service occupations; trades, transport and equipment operators and related occupations; natural resources, agriculture, and related production occupations; and occupations in manufacturing and utilities. Census counts for people are randomly rounded up or down to the nearest number divisible by 5, which causes some minor imprecision.

**^g^**Range of persons per dwelling.

^h^Percentage of people in the area who self-identified as a visible minority. Census counts for people are randomly rounded up or down to the nearest number divisible by 5, which causes some minor imprecision.

**Supplementary Table 2.** Characteristics of subjects tested for SARS-CoV-2 with COVID-19-relevant symptoms during the period November 22 to December 26, 2021, comparing Omicron cases and SARS-CoV-2-negative controls by time period

|  | **SARS-CoV-2 negative,**  **22 Nov to**  **12 Dec 2021,**  **n (%)^a^** | **Omicron,**  **22 Nov to**  **12 Dec 2021,**  **n (%)^a^** | **SD^b^** | **SARS-CoV-2 negative,**  **13 Dec to**  **19 Dec 2021,**  **n (%)^a^** | **Omicron,**  **13 Dec to**  **19 Dec 2021,**  **n (%)^a^** | **SD^b^** | **SARS-CoV-2 negative,**  **20 Dec to**  **26 Dec 2021,**  **n (%)^a^** | **Omicron,**  **20 Dec to**  **26 Dec 2021,**  **n (%)^a^** | **SD^b^** |
| --- | --- | --- | --- | --- | --- | --- | --- | --- | --- |
| **Total** | N=83,111 | N=776 |  | N=41,090 | N=5,049 |  | N=41,894 | N=10,309 |  |
| Subject characteristics |  |  |  |  |  |  |  |  |  |
| Age (years), mean (standard deviation) | 41.9 ± 16.5 | 32.2 ± 14.0 | 0.63 | 42.0 ± 16.4 | 36.2 ± 13.8 | 0.39 | 42.1 ± 16.7 | 36.3 ± 14.2 | 0.38 |
| Age group (years) |  |  |  |  |  |  |  |  |  |
| 18–29 | 22,039 (26.5%) | 424 (54.6%) | 0.60 | 10,941 (26.6%) | 2,071 (41.0%) | 0.31 | 11,820 (28.2%) | 4,341 (42.1%) | 0.29 |
| 30–39 | 21,572 (26.0%) | 135 (17.4%) | 0.21 | 10,308 (25.1%) | 1,141 (22.6%) | 0.06 | 9,999 (23.9%) | 2,201 (21.4%) | 0.06 |
| 40–49 | 14,239 (17.1%) | 114 (14.7%) | 0.07 | 7,171 (17.5%) | 948 (18.8%) | 0.03 | 6,473 (15.5%) | 1,767 (17.1%) | 0.05 |
| 50–59 | 11,676 (14.0%) | 71 (9.1%) | 0.15 | 5,879 (14.3%) | 573 (11.3%) | 0.09 | 6,429 (15.3%) | 1,282 (12.4%) | 0.08 |
| 60–69 | 7,789 (9.4%) | 17 (2.2%) | 0.31 | 3,995 (9.7%) | 215 (4.3%) | 0.22 | 4,187 (10.0%) | 492 (4.8%) | 0.20 |
| 70–79 | 3,827 (4.6%) | 10-14 (1.3-1.8%) | 0.16-0.20 | 1,922 (4.7%) | 64 (1.3%) | 0.20 | 2,018 (4.8%) | 160 (1.6%) | 0.19 |
| ≥80 | 1,969 (2.4%) | ≤5 (≤0.6%) | 0.14-0.20 | 874 (2.1%) | 37 (0.7%) | 0.12 | 968 (2.3%) | 66 (0.6%) | 0.14 |
| Male sex | 33,648 (40.5%) | 348 (44.8%) | 0.09 | 16,779 (40.8%) | 2,553 (50.6%) | 0.20 | 16,769 (40.0%) | 4,956 (48.1%) | 0.16 |
| Any comorbidity^c^ | 36,661 (44.1%) | 264 (34.0%) | 0.21 | 17,565 (42.7%) | 1,852 (36.7%) | 0.12 | 18,105 (43.2%) | 3,744 (36.3%) | 0.14 |
| Number of SARS-CoV-2 tests within 3 months prior to 14 Dec 2020 |  |  |  |  |  |  |  |  |  |
| 0 | 60,255 (72.5%) | 558 (71.9%) | 0.01 | 29,981 (73.0%) | 3,880 (76.8%) | 0.09 | 30,847 (73.6%) | 8,043 (78.0%) | 0.10 |
| 1 | 16,139 (19.4%) | 158 (20.4%) | 0.02 | 7,972 (19.4%) | 912 (18.1%) | 0.03 | 7,883 (18.8%) | 1,720 (16.7%) | 0.06 |
| ≥2 | 6,717 (8.1%) | 60 (7.7%) | 0.01 | 3,137 (7.6%) | 257 (5.1%) | 0.10 | 3,164 (7.6%) | 546 (5.3%) | 0.09 |
| Receipt of 2019-2020 and/or 2020-2021 influenza vaccination | 28,656 (34.5%) | 195 (25.1%) | 0.21 | 14,832 (36.1%) | 1,312 (26.0%) | 0.22 | 14,942 (35.7%) | 2,516 (24.4%) | 0.25 |
| Public health unit region^d^ |  |  |  |  |  |  |  |  |  |
| Central East | 7,677 (9.2%) | 25 (3.2%) | 0.25 | 3,473 (8.5%) | 275 (5.4%) | 0.12 | 2,904 (6.9%) | 532 (5.2%) | 0.07 |
| Central West | 14,860 (17.9%) | 156 (20.1%) | 0.06 | 7,905 (19.2%) | 1,149 (22.8%) | 0.09 | 8,004 (19.1%) | 2,605 (25.3%) | 0.15 |
| Durham | 5,980 (7.2%) | 44 (5.7%) | 0.06 | 2,688 (6.5%) | 337 (6.7%) | 0.01 | 2,647 (6.3%) | 670 (6.5%) | 0.01 |
| Eastern | 5,524 (6.6%) | 151 (19.5%) | 0.39 | 2,747 (6.7%) | 317 (6.3%) | 0.02 | 2,185 (5.2%) | 405 (3.9%) | 0.06 |
| North | 6,529 (7.9%) | <=5 (0.6%) | 0.36 | 2,530 (6.2%) | 64 (1.3%) | 0.26 | 2,769 (6.6%) | 199 (1.9%) | 0.23 |
| Ottawa | 3,221 (3.9%) | 78 (10.1%) | 0.24 | 1,974 (4.8%) | 380 (7.5%) | 0.11 | 2,365 (5.6%) | 527 (5.1%) | 0.02 |
| Peel | 9,363 (11.3%) | 86 (11.1%) | 0.01 | 4,439 (10.8%) | 740 (14.7%) | 0.12 | 4,733 (11.3%) | 1,728 (16.8%) | 0.16 |
| South West | 9,103 (11.0%) | 28 (3.6%) | 0.29 | 4,568 (11.1%) | 201 (4.0%) | 0.27 | 4,677 (11.2%) | 770 (7.5%) | 0.13 |
| Toronto | 14,433 (17.4%) | 148 (19.1%) | 0.04 | 7,778 (18.9%) | 1,143 (22.6%) | 0.09 | 8,301 (19.8%) | 1,820 (17.7%) | 0.06 |
| York | 6,069 (7.3%) | 51 (6.6%) | 0.03 | 2,810 (6.8%) | 413 (8.2%) | 0.05 | 3,158 (7.5%) | 1,012 (9.8%) | 0.08 |
| Household income quintile^d, e^ |  |  |  |  |  |  |  |  |  |
| 1 (lowest) | 13,798 (16.6%) | 71 (9.1%) | 0.22 | 6,255 (15.2%) | 605 (12.0%) | 0.09 | 6,248 (14.9%) | 1,239 (12.0%) | 0.08 |
| 2 | 15,485 (18.6%) | 105 (13.5%) | 0.14 | 7,531 (18.3%) | 787 (15.6%) | 0.07 | 7,574 (18.1%) | 1,643 (15.9%) | 0.06 |
| 3 | 16,421 (19.8%) | 125 (16.1%) | 0.10 | 8,007 (19.5%) | 914 (18.1%) | 0.04 | 8,428 (20.1%) | 2,056 (19.9%) | 0 |
| 4 | 17,977 (21.6%) | 194 (25.0%) | 0.08 | 8,806 (21.4%) | 1,139 (22.6%) | 0.03 | 9,302 (22.2%) | 2,456 (23.8%) | 0.04 |
| 5 (highest) | 19,016 (22.9%) | 275 (35.4%) | 0.28 | 10,301 (25.1%) | 1,572 (31.1%) | 0.14 | 10,170 (24.3%) | 2,871 (27.8%) | 0.08 |
| Essential workers quintile^d, f^ |  |  |  |  |  |  |  |  |  |
| 1 (0%–32.5%) | 17,484 (21.0%) | 286 (36.9%) | 0.35 | 9,811 (23.9%) | 1,585 (31.4%) | 0.17 | 10,231 (24.4%) | 2,684 (26.0%) | 0.04 |
| 2 (32.5%–42.3%) | 19,900 (23.9%) | 252 (32.5%) | 0.19 | 10,140 (24.7%) | 1,299 (25.7%) | 0.02 | 10,398 (24.8%) | 2,952 (28.6%) | 0.09 |
| 3 (42.3%–49.8%) | 16,962 (20.4%) | 111 (14.3%) | 0.16 | 7,978 (19.4%) | 953 (18.9%) | 0.01 | 8,478 (20.2%) | 2,001 (19.4%) | 0.02 |
| 4 (50.0%–57.5%) | 15,110 (18.2%) | 80 (10.3%) | 0.23 | 7,200 (17.5%) | 696 (13.8%) | 0.10 | 6,865 (16.4%) | 1,500 (14.6%) | 0.05 |
| 5 (57.5%–100%) | 12,986 (15.6%) | 41 (5.3%) | 0.34 | 5,702 (13.9%) | 479 (9.5%) | 0.14 | 5,678 (13.6%) | 1,104 (10.7%) | 0.09 |
| Persons per dwelling quintile^d,^ **^g^** |  |  |  |  |  |  |  |  |  |
| 1 (0–2.1) | 15,056 (18.1%) | 110 (14.2%) | 0.11 | 7,726 (18.8%) | 790 (15.6%) | 0.08 | 7,833 (18.7%) | 1,630 (15.8%) | 0.08 |
| 2 (2.2–2.4) | 14,188 (17.1%) | 115 (14.8%) | 0.06 | 6,910 (16.8%) | 587 (11.6%) | 0.15 | 6,978 (16.7%) | 1,244 (12.1%) | 0.13 |
| 3 (2.5–2.6) | 11,086 (13.3%) | 97 (12.5%) | 0.03 | 5,424 (13.2%) | 631 (12.5%) | 0.02 | 5,552 (13.3%) | 1,196 (11.6%) | 0.05 |
| 4 (2.7–3.0) | 20,230 (24.3%) | 199 (25.6%) | 0.03 | 10,152 (24.7%) | 1,364 (27.0%) | 0.05 | 10,111 (24.1%) | 2,589 (25.1%) | 0.02 |
| 5 (3.1–5.7) | 21,849 (26.3%) | 247 (31.8%) | 0.12 | 10,605 (25.8%) | 1,637 (32.4%) | 0.15 | 11,155 (26.6%) | 3,573 (34.7%) | 0.17 |
| Self-identified visible minority quintile^d, h^ |  |  |  |  |  |  |  |  |  |
| 1 (0.0%–2.2%) | 13,212 (15.9%) | 92 (11.9%) | 0.12 | 6,166 (15.0%) | 416 (8.2%) | 0.21 | 5,992 (14.3%) | 937 (9.1%) | 0.16 |
| 2 (2.2%–7.5%) | 15,331 (18.4%) | 116 (14.9%) | 0.09 | 7,418 (18.1%) | 709 (14.0%) | 0.11 | 7,047 (16.8%) | 1,420 (13.8%) | 0.08 |
| 3 (7.5%–18.7%) | 16,103 (19.4%) | 201 (25.9%) | 0.16 | 8,337 (20.3%) | 1,091 (21.6%) | 0.03 | 8,142 (19.4%) | 2,123 (20.6%) | 0.03 |
| 4 (18.7%–43.5%) | 17,816 (21.4%) | 190 (24.5%) | 0.07 | 9,287 (22.6%) | 1,392 (27.6%) | 0.11 | 10,013 (23.9%) | 2,680 (26.0%) | 0.05 |
| 5 (43.5%–100%) | 19,984 (24.0%) | 171 (22.0%) | 0.05 | 9,623 (23.4%) | 1,404 (27.8%) | 0.10 | 10,457 (25.0%) | 3,081 (29.9%) | 0.11 |
| Prior positive SARS-CoV-2 test | 3,418 (4.1%) | 9 (1.2%) | 0.19 | 1,462 (3.6%) | 42 (0.8%) | 0.19 | 1,520 (3.6%) | 66 (0.6%) | 0.21 |
| COVID-19 vaccine characteristics |  |  |  |  |  |  |  |  |  |
| Unvaccinated | 4,453 (5.4%) | 35 (4.5%) | 0.04 | 1,605 (3.9%) | 236 (4.7%) | 0.04 | 1,524 (3.6%) | 526 (5.1%) | 0.07 |
| Received 2-dose primary series only (with at least 1 mRNA vaccine) | 73,350 (88.3%) | 722 (93.0%) | 0.16 | 34,255 (83.4%) | 4,525 (89.6%) | 0.18 | 30,019 (71.7%) | 8,605 (83.5%) | 0.29 |
| Received BNT162b2 for third dose | 4,524 (5.4%) | 14-18 (1.8-2.3%) | 0.16-0.20 | 4,403 (10.7%) | 255 (5.1%) | 0.21 | 8,209 (19.6%) | 953 (9.2%) | 0.30 |
| Received mRNA-1273 for third dose | 784 (0.9%) | ≤5 (≤0.6%) | 0.03-0.11 | 827 (2.0%) | 33 (0.7%) | 0.12 | 2,142 (5.1%) | 225 (2.2%) | 0.16 |
| Time since second dose |  |  |  |  |  |  |  |  |  |
| 7-59 days | 2,631 (3.2%) | 16 (2.1%) | 0.07 | 815 (2.0%) | 75 (1.5%) | 0.04 | 640 (1.5%) | 141 (1.4%) | 0.01 |
| 60-119 days | 7,063 (8.5%) | 49 (6.3%) | 0.08 | 2,394 (5.8%) | 313 (6.2%) | 0.02 | 2,085 (5.0%) | 645 (6.3%) | 0.06 |
| 120-179 days | 56,227 (67.7%) | 561 (72.3%) | 0.10 | 23,162 (56.4%) | 3,036 (60.1%) | 0.08 | 17,369 (41.5%) | 4,975 (48.3%) | 0.14 |
| 180-239 days | 5,810 (7.0%) | 81 (10.4%) | 0.12 | 7,242 (17.6%) | 1,044 (20.7%) | 0.08 | 9,380 (22.4%) | 2,695 (26.1%) | 0.09 |
| ≥240 days | 1,619 (1.9%) | 15 (1.9%) | 0 | 642 (1.6%) | 57 (1.1%) | 0.04 | 545 (1.3%) | 149 (1.4%) | 0.01 |
| Time since third dose |  |  |  |  |  |  |  |  |  |
| No third dose (i.e., only 2 doses) | 73,350 (88.3%) | 722 (93.0%) | 0.16 | 34,255 (83.4%) | 4,525 (89.6%) | 0.18 | 30,019 (71.7%) | 8,605 (83.5%) | 0.29 |
| 0-6 days | 1,040 (1.3%) | 6 (0.8%) | 0.05 | 1,405 (3.4%) | 127 (2.5%) | 0.05 | 4,127 (9.9%) | 672 (6.5%) | 0.12 |
| 7-59 days | 3,845 (4.6%) | 8-12 (1.0-1.5%) | 0.18-0.22 | 3,528 (8.6%) | 152 (3.0%) | 0.24 | 5,844 (13.9%) | 474 (4.6%) | 0.33 |
| ≥60 days | 423 (0.5%) | ≤5 (0.1%) | 0.02-0.07 | 297 (0.7%) | 9 (0.2%) | 0.08 | 380 (0.9%) | 32 (0.3%) | 0.08 |
| Interval between first dose and second dose |  |  |  |  |  |  |  |  |  |
| 15-34 days | 11,776 (14.2%) | 159 (20.5%) | 0.17 | 5,538 (13.5%) | 705 (14.0%) | 0.01 | 5,591 (13.3%) | 1,418 (13.8%) | 0.01 |
| 35-55 days | 26,518 (31.9%) | 323 (41.6%) | 0.20 | 13,760 (33.5%) | 1,978 (39.2%) | 0.12 | 14,167 (33.8%) | 3,871 (37.5%) | 0.08 |
| ≥56 days | 40,364 (48.6%) | 259 (33.4%) | 0.31 | 20,187 (49.1%) | 2,130 (42.2%) | 0.14 | 20,612 (49.2%) | 4,494 (43.6%) | 0.11 |
| Interval between second dose and third dose |  |  |  |  |  |  |  |  |  |
| No third dose (i.e., only 2 doses) | 73,350 (88.3%) | 722 (93.0%) | 0.16 | 34,255 (83.4%) | 4,525 (89.6%) | 0.18 | 30,019 (71.7%) | 8,605 (83.5%) | 0.29 |
| ≤111 days | 177 (0.2%) | ≤5 (0.1%) | 0.02 | 83 (0.2%) | 7 (0.1%) | 0.02 | 88 (0.2%) | 10 (0.1%) | 0.03 |
| 112-167 days | 690 (0.8%) | ≤5 (0.1%) | 0.10 | 475 (1.2%) | 10 (0.2%) | 0.12 | 853 (2.0%) | 111 (1.1%) | 0.08 |
| ≥168 days | 4,441 (5.3%) | 14-18 (1.8-2.3%) | 0.15-0.19 | 4,672 (11.4%) | 271 (5.4%) | 0.22 | 9,410 (22.5%) | 1,057 (10.3%) | 0.33 |

**^a^**Proportion reported, unless stated otherwise.

**^b^**SD=standardized difference. Standardized differences of >0.10 are considered clinically relevant. Comparison of Omicron-positive cases with SARS-CoV-2-negative controls in each time period.

^c^Comorbidities include chronic respiratory diseases, chronic heart diseases, hypertension, diabetes, immunocompromising conditions due to underlying diseases or therapy, autoimmune diseases, chronic kidney disease, advanced liver disease, dementia/frailty and history of stroke or transient ischemic attack.

^d^The sum of counts does not equal the column total because of individuals with missing information (<1.0%) for this characteristic.

^e^Household income quintile has variable cut-off values in each city/Census area to account for cost of living. A dissemination area (DA) being in quintile 1 means it is among the lowest 20% of DAs in its city by income.

^f^Percentage of people in the area working in the following occupations: sales and service occupations; trades, transport and equipment operators and related occupations; natural resources, agriculture, and related production occupations; and occupations in manufacturing and utilities. Census counts for people are randomly rounded up or down to the nearest number divisible by 5, which causes some minor imprecision.

**^g^**Range of persons per dwelling.

^h^Percentage of people in the area who self-identified as a visible minority. Census counts for people are randomly rounded up or down to the nearest number divisible by 5, which causes some minor imprecision.

**Supplementary Table 3.** Number of Omicron and Delta severe outcomes (hospitalization or death) by laboratory testing categorization during the period December 6 to 26, 2021

| **Classification criteria** | **6 Dec to 12 Dec 2021** | **13 Dec to 19 Dec 2021** | **20 Dec to 26 Dec 2021** |
| --- | --- | --- | --- |
| Omicron confirmed via WGS or SGTF | 7 | 28 | 53 |
| Omicron classified if collection date ≥ 21 December 2021 and no or inconclusive SGTF results | n/a | n/a | 177 |
| Delta confirmed via WGS or SGTF | 121 | 115 | 61 |
| No classification (excluded) | 62 | 65 | 15 |

WGS: whole genome sequencing; SGTF: S-gene target failure

**Supplementary Table 4.** Estimates of vaccine effectiveness* against severe outcomes (hospitalization or death) during the period December 6 to 26, 2021 using the less specific definition of Omicron and Delta

| **Outcome** | **Doses** | **Vaccine products** | **Days since latest dose** | **Vaccine effectiveness against Omicron (95% CI)** |
| --- | --- | --- | --- | --- |
| Severe outcomes | First 2 doses | ≥1 mRNA vaccine | 7-59 | 89 (64, 97) |
|  |  |  | 60-119 | 85 (69, 92) |
|  |  |  | 120-179 | 92 (89, 95) |
|  |  |  | 180-239 | 93 (89, 95) |
|  |  |  | ≥240 | 93 (77, 98) |
|  | Third dose | Any mRNA vaccine | 0-6 | 94 (90, 96) |
|  |  |  | ≥7 | 98 (97, 99) |
|  |  | BNT162b2 | 0-6 | 95 (91, 97) |
|  |  |  | ≥7 | 98 (97, 99) |
|  |  | mRNA-1273 | 0-6 | 91 (81, 96) |
|  |  |  | ≥7 | 98 (96, 99) |

*Vaccine effectiveness estimates adjusted for: age (in 10-year age bands), sex, public health unit region of residence, number of SARS-CoV-2 PCR tests during the 3 months prior to December 14, 2020, past SARS-CoV-2 infection >90 days prior to index date, comorbidities, influenza vaccination status during the 2019/2020 and/or 2020/2021 influenza seasons, and neighbourhood-level information on median household income, proportion of the working population employed as non-health essential workers, mean number of persons per dwelling, and proportion of the population who self-identify as a visible minority.

**Supplementary Figure 1.** Estimates of vaccine effectiveness against any infection (irrespective of symptoms or severity) with Omicron or Delta, comparing results reported in an initial pre-print using an earlier time period (November 22 to December 19, 2021; panel A) to an updated time period (December 6 to 26, 2021; panel B). The second period excluded the first two weeks (with only 47 Omicron cases) and added an additional week. Panel A included individuals with whole genome sequencing (WGS) or S-gene target failure (SGTF) results, as well as those with missing or inconclusive SGTF results prior to December 3, 2021 (when the prevalence of Omicron was <5% in the province), whereas panel B was restricted to WGS or SGTF results. Biases introduced by differential access or test-seeking behaviour by exposure or vaccination status persisted over time when using any infection as the outcome, reinforcing the importance of using symptomatic infection as a more appropriate outcome for estimating vaccine effectiveness.

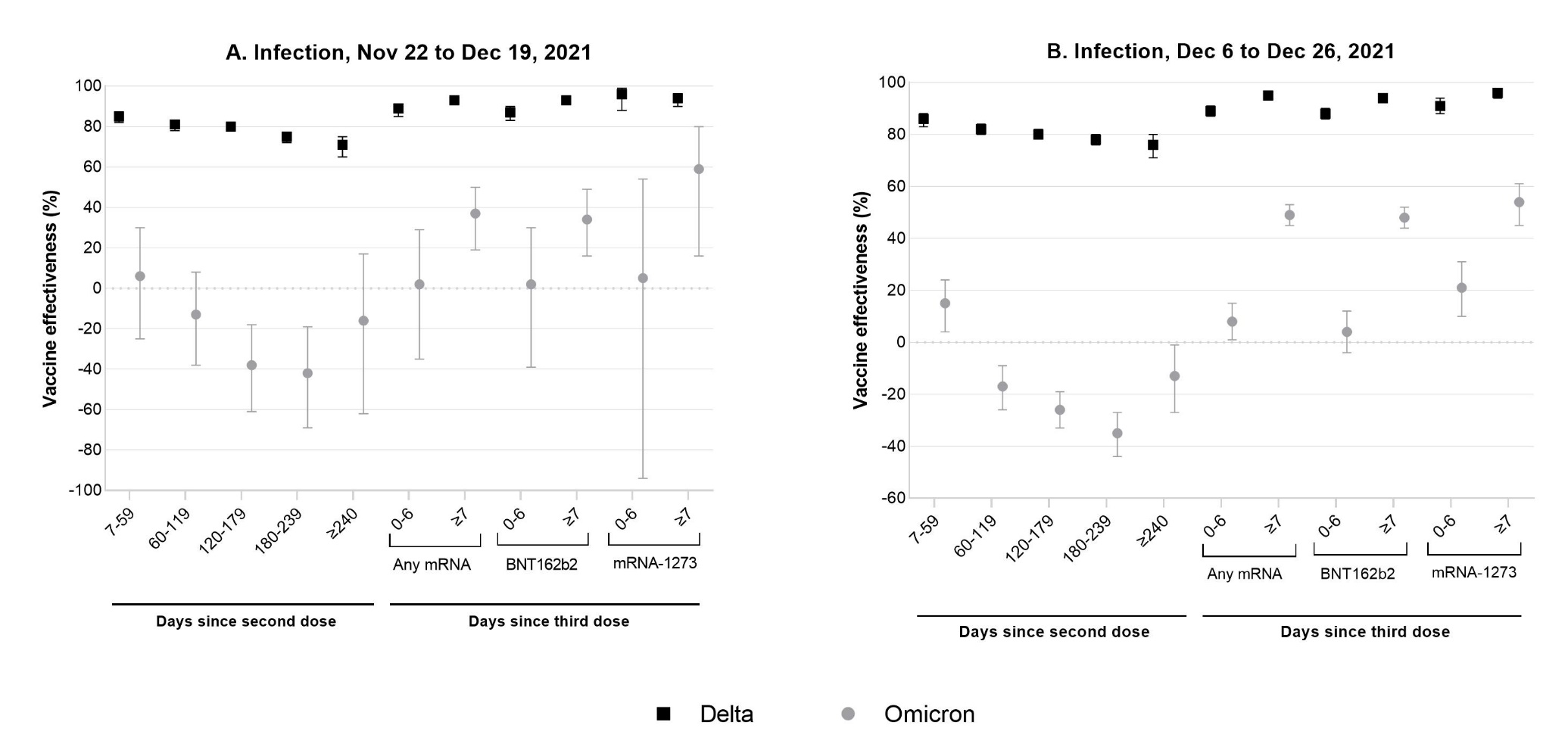
